# Incidence of Glomerulonephritis after SARS-CoV-2 mRNA Vaccination

**DOI:** 10.1101/2022.05.18.22275112

**Authors:** Matthias Diebold, Eleonore Locher, Philipp Boide, Annette Enzler-Tschudy, Anna Faivre, Ingeborg Fischer, Birgit Helmchen, Helmut Hopfer, Min Jeong Kim, Solange Moll, Giliane Nanchen, Samuel Rotman, Charalampos Saganas, Harald Seeger, Andreas D. Kistler

**Author notes:** **Corresponding Author:** PD Dr. med. Andreas D. Kistler, Department of Medicine, Cantonal Hospital Frauenfeld, Pfaffenholzstrasse 4, 8501 Frauenfeld.

## Abstract

Numerous cases of glomerulonephritis manifesting shortly after SARS-CoV-2 vaccination have been reported, but causality remains unproven. We studied the association between mRNA-based SARS-CoV-2 vaccination and new-onset glomerulonephritis using a nationwide retrospective cohort and case-cohort design. Data from all Swiss pathology institutes processing native kidney biopsies served to calculate incidence of IgA nephropathy, pauci-immune necrotizing glomerulonephritis, minimal change disease and membranous nephropathy. The observed incidence during the vaccination campaign (Jan to Aug 2021) was not different from the expected incidence based on the years 2015 to 2019 (incidence rate ratio 0.86, 95%-credible interval 0.73–1.02) and did not cross the upper boundary of the 95% credible interval for any month. Among 111 patients aged >18 years with newly diagnosed glomerulonephritis between January and August 2021, 38.7% had received at least one vaccine dose before biopsy, compared to 39.5% of the general Swiss population matched for age and calendar-time. The estimated risk ratio for the development of new-onset biopsy-proven glomerulonephritis was 0.97 (95% CI 0.66–1.42, *P*=0.95) in vaccinated vs. unvaccinated individuals. Patients with glomerulonephritis manifesting within 4 weeks after vaccine did not differ clinically from the rest of the cohort. Results were consistent across all types of glomerulonephritis with the possible exception of minimal change disease. In conclusion, vaccination against SARS-CoV-2 was not associated with new-onset glomerulonephritis in these two complementary studies. Most temporal associations between SARS-CoV-2 vaccination and glomerulonephritis are likely coincidental.

## Introduction

Following emergency use authorization for the first vaccines developed against SARS-CoV-2 in many countries in late 2020, an unprecedented vaccination-campaign was initiated worldwide in 2021. In Europe and North America, the majority of people received mRNA-based vaccines, mostly either BNT162b2 (Pfizer-BioNTech) or mRNA-1273 (Moderna). Both vaccines had been tested in large randomized controlled trials with excellent safety profiles.^1, 2^ However, these registration studies were not powered to detect rare vaccine side effects. During the worldwide vaccination-campaign numerous rare adverse events with a temporal association to SARS-CoV-2 vaccination were observed,^3^ including various autoimmune phenomena.^4^ Among such associations, many new-onset and recurrent cases of glomerulonephritis have been reported.^5-11^ Specific types of glomerulonephritis most often reported after SARS-CoV-2 vaccination include IgA nephropathy (IgAN), minimal change disease (MCD), membranous nephropathy (MN) and pauci-immune necrotizing glomerulonephritis (PINGN), the renal manifestation of antineutrophil cytoplasmic antibody (ANCA)–associated vasculitis (AAV).

While mechanistically plausible,^5, 7, 11^ a causal link between SARS-CoV-2 vaccination and glomerulonephritis has not been formally established. Published case series lack control groups, and estimates of incidence of these glomerulonephritides before and during the vaccination campaign have not been published yet. Given the high number of vaccine doses administered, many cases of new onset glomerulonephritis would be expected to occur in temporal proximity to vaccination by pure coincidence.

Here, we tested the hypothesis that SARS-CoV-2 mRNA vaccines increase the incidence of several types of glomerulonephritis against the null hypothesis that reported temporal associations can be attributed to a by-chance-effect. We compared the observed incidence of glomerulonephritis in Switzerland during the vaccination campaign in 2021 to the expected incidence based on a baseline period (2015 to 2019). We further compared the vaccination history of patients with glomerulonephritis newly diagnosed during the vaccination campaign to the general population matched for age and timepoint and we characterized patients with new-onset glomerulonephritis in temporal association with vaccination.

## Methods

### Study design

We performed two complementary, interconnected studies. For the first study, all Swiss pathology institutes analyzing kidney biopsies provided biopsy date, age, sex and histological diagnosis for all patients aged >18 years with histological diagnosis of IgAN, PINGN, MCD or MN from January 1, 2015 through August 31, 2021. These data, together with census data for each year (2015 – 2021) on the Swiss population aged >18 years, were used to calculate the incidence of all four types of glomerulonephritis. For the second study, all of the above-mentioned patients with biopsy date between January 1 and August 31, 2021, were eligible. Clinical information was gathered and vaccination history compared to the general Swiss population, matched for age and time-point during the vaccination campaign.

Both studies were approved by the Ethics Committee of Eastern Switzerland. The first study was exempt from patient consent, because only minimal data were included and it would have been impossible to obtain consent from all patients, while inclusion of all cases was essential to calculate true incidence. For the second study, written informed consent was obtained from all participants, and the study was conducted in accordance with Good Clinical Practice guidelines and the principles of the Declaration of Helsinki.^12^ We followed the STROBE Guidelines for reporting observational studies.^13^

### Data collection

For the first study, local nephropathologists extracted relevant data from an existing database or performed a search of their electronic records. We limited our study to native kidney biopsies of persons aged >18 years with IgAN, PINGN, MCD and MN. These glomerulonephritides belong to the most frequent that can be unambiguously diagnosed by histology and have been reported after SARS-CoV-2 vaccination. Lupus nephritis was not included, because repeat biopsies are often performed for this glomerulonephritis, constituting a significant proportion of all lupus nephritis biopsies. We did not include focal segmental glomerulosclerosis (FSGS), since most FSGS cases are secondary, caused by distinct pathophysiological mechanisms and not reliably distinguishable from primary FSGS by histology.^14^ MN was included in the analysis, because secondary MN constitutes a minority of cases,^15^ but lupus nephritis class V was excluded for reasons outlined above. As information on prior kidney biopsy was not available from the records of some pathology institutes and because repeat biopsies constitute only a small fraction of all biopsies, we included both, new diagnoses and repeat biopsies in the first study.

For the second study, nephropathologists provided the referring nephrologists’ names for all eligible patients (>18 years old with histological diagnosis of IgAN, PINCG, MCD or MN between January 1 and August 31, 2021) to the investigators. We contacted all nephrology divisions and practices who had sent in biopsy specimens of eligible patients providing age, sex, histological diagnosis and date of biopsy to help them identify their patients and to contact them for study participation. Clinical and epidemiological data were collected in a case report form (CRF) completed by the treating nephrologist and in a questionnaire completed by the patient. All data were reviewed and the time-point of symptom onset or laboratory abnormalities attributable to the glomerular disease was adjudicated by the study investigators (see Supplementary Methods).

Pathologists provided complete data between September and December 2021, nephrologists were contacted between October and December 2021 and all questionnaires and CRF returned by February 22, 2022, were included in the analysis.

Data on the types and numbers of vaccine doses by calendar week and age decade for the general population were downloaded from the Swiss Federal Office of Public Health (https://www.covid19.admin.ch/de/vaccination/doses, accessed on January 31, 2022).

### Statistical analysis

For the first study, a Bayesian Poisson regression model was used to predict the expected incidence of glomerulonephritis cases for each month in 2021 based on data for the years 2015 to 2019, excluding 2020 because we expected underdiagnosis early in the pandemic due to lock-down measures. We calculated incidence rate ratios for the total of all four glomerulonephritides and for each diagnosis separately by dividing the observed cases in 2021 by the expected cases and expressed uncertainty using 95% credible intervals. Because the availability of serologic testing for anti-PLA2R autoantibodies may have influenced biopsy practice over time,^16^ we performed a sensitivity analysis excluding MN.

For the second study, we chose a control group from the general population by determining for every patient the percentage of individuals from the general population of the same age decade (the decade 20-29 was used for the few patients aged 18 or 19 years) who had received a first, second, or any vaccine dose in every one-week interval prior to (a) the calendar week of the biopsy date and (b) the calendar week of onset of symptoms or detection of laboratory abnormalities attributable to the glomerulonephritis and used 1000 controls per patient. For the comparison between controls and patients we used risk ratios by unconditional maximum likelihood estimation given that the total number of vaccinated and unvaccinated persons in Switzerland (including both, cases and non-cases) was available and assuming that a representative sample of all patients with new-onset glomerulonephritis was included in the second study. Discrete variables are expressed as counts (percentage) and continuous variables as median and interquartile range [IQR]. Comparisons between groups were made using Kruskal-Wallis’ test and Pearson’s X^2^ test, as appropriate.

Statistical analyses were performed using R software, version R version 4.0.2 (R Core Team 2020. R: A language and environment for statistical computing. R Foundation for Statistical Computing, Vienna, Austria. URL https://www.R-project.org/). A list of the packages used for this analysis is provided in the Supplementary Methods.

## Results

### Incidence of glomerulonephritis before and during the vaccination campaign

During the baseline period (2015 to 2019), the incidence of IgAN, PINGN, MCD and MN was 23.8, 11.9, 5.1 and 9.3 cases per million population per year, respectively and remained stable over time (Supplementary Figure 1). In 2020, we found a reduced incidence during the first infection wave and the consecutive public lock-down measures, which was attributable to IgAN (Supplementary Figure 2). The first vaccines were administered in Switzerland in mid-December 2020, the number of vaccinations increased to reach a maximum in June 2021, and declined thereafter (**Figure 1, upper left panel**). The observed incidence of glomerulonephritis from January to August 2021 compared to the expected incidence is shown in **Figure 1**. The observed incidence was compatible with the expected incidence for all months during the vaccination campaign (**Table 1**) with an overall incidence rate ratio of 0.86 (95%-credible interval 0.73 to 1.02) for the entire study period (January to August 2021). In an additional analysis comparing the weekly incidence during the entire study period or the peak of the vaccination campaign (May to August 2021) to the corresponding calendar time of the baseline period, we also did not find any difference (Supplementary Table 1).

**Fig 1.**
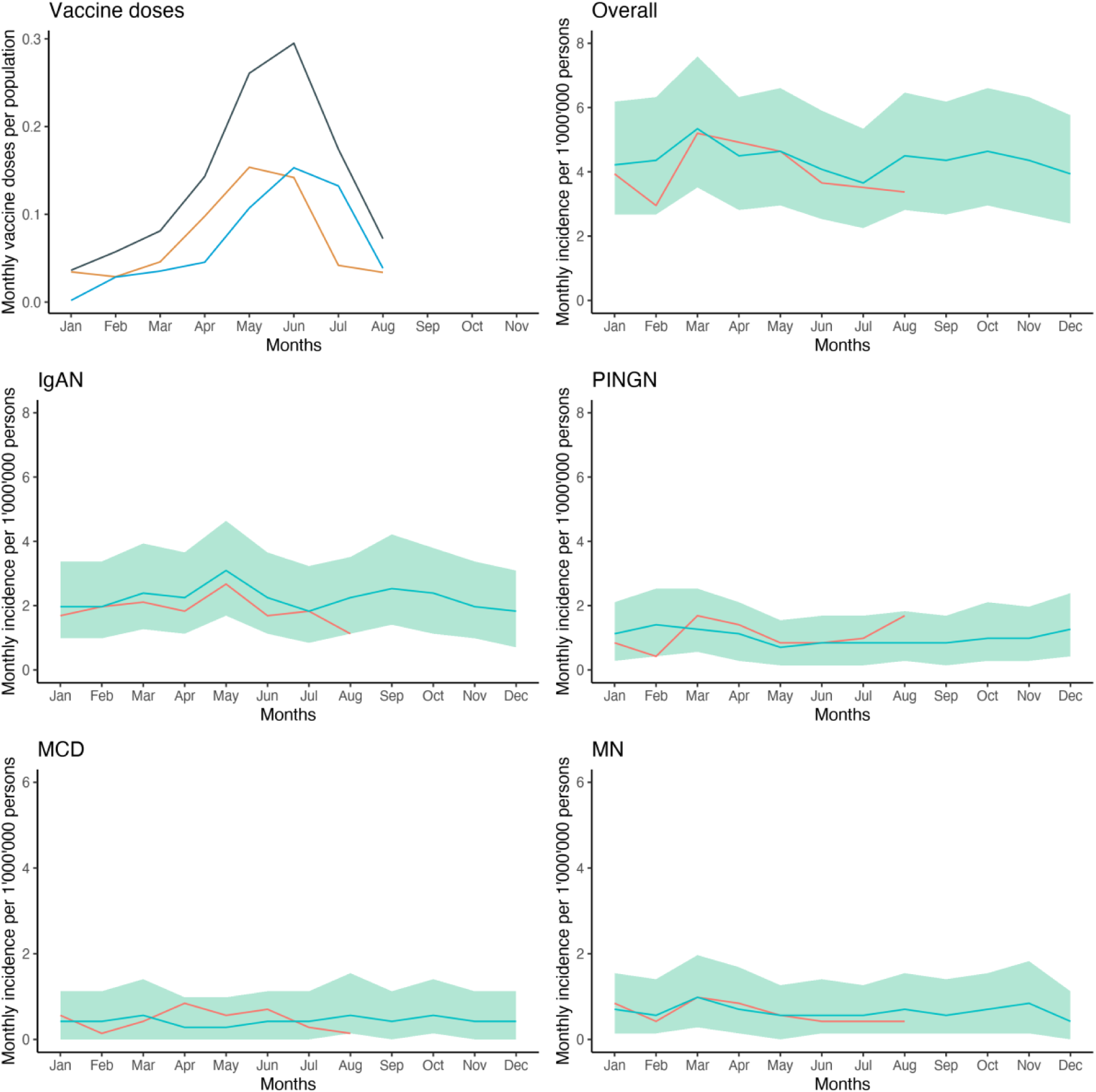
Expected and observed incidence of glomerulonephritis during the vaccination campaign. Shown is the number of first (orange), second (blue) and total (grey) doses of SARS-CoV-2 vaccines as a fraction of all patients aged ≥ 20 years (upper left panel) and the observed incidence of glomerulonephritis in patients aged ≥ 18 years from January to August 2021 (red line) compared to the expected incidence based on the years 2015 to 2019 (blue line) with 95% credible intervals (green shading) for the sum of all four glomerulonephritides, and by diagnosis.

**Table 1.** Observed monthly incidence of glomerulonephritis during the vaccination campaign compared to the baseline period (2015 to 2019) and the expected incidence, relative risk ratio and number of vaccine doses administered in the corresponding month.

|  | Jan | Feb | Mar | Apr | May | Jun | Jul | Aug |
| --- | --- | --- | --- | --- | --- | --- | --- | --- |
| Incidence 2021 | 3.94 | 2.95 | 5.2 | 4.92 | 4.64 | 3.65 | 3.51 | 3.37 |
| Observed incidence 2015-2019 | 4.05 | 4.18 | 5.11 | 4.29 | 4.37 | 3.92 | 3.51 | 4.26 |
| Expected incidence (95% credible intervals) | 4.22<br>(2.67 to 6.04) | 4.36<br>(2.67 to 6.32) | 5.34<br>(3.51 to 7.45) | 4.5<br>(2.81 to 6.47) | 4.5<br>(2.81 to 6.47) | 4.08<br>(2.53 to 5.90) | 3.65<br>(2.25 to 5.90) | 4.36<br>(2.81 to 6.47) |
| Relative incidence rate ratio (95% credible intervals) | 0.93<br>(0.64 to 1.47) | 0.68<br>(0.47 to 1.11) | 0.97<br>(0.69 to 1.48) | 1.09<br>(0.76 to 1.75) | 1.03<br>(0.70 to 1.65) | 0.90<br>(0.62 to 1.44) | 0.96<br>(0.66 to 1.56) | 0.77<br>(0.52 to 1.20) |
| Vaccine doses per population | 0.04 | 0.06 | 0.08 | 0.14 | 0.26 | 0.29 | 0.17 | 0.07 |

Incidence is given as the number of biopsy-proven glomerulonephritis cases (IgAN, PIGN, MCD and MN) per million population aged >18 years. Vaccine doses per population are given as doses per population. The expected incidence was calculated using a Bayesian model based on each month for the years 2015 to 2019. The relative incidence rate denotes the ratio between observed and expected cases.

### Characteristics of patients with new-onset glomerulonephritis during the vaccination campaign

Overall, 229 patients aged >18 years had a biopsy-proven diagnosis of IgAN (n = 106), PINGN (n = 62), MCD (n = 26) or MN (n = 35) between January and August 2021 in Switzerland. 125 (54.6%) of these patients could be included in the second study. Reasons for non-inclusion are shown in the study flowchart (**Figure 2**). 14 patients (11.2%) with repeat biopsies of a previously established diagnosis were not included in the analysis, and their clinical characteristics and reasons for repeat biopsy are shown in Supplementary Table 2. Clinical characteristics of all patients with a new diagnosis of glomerulonephritis (n = 111) are shown in **Table 2**.

**Fig 2.**
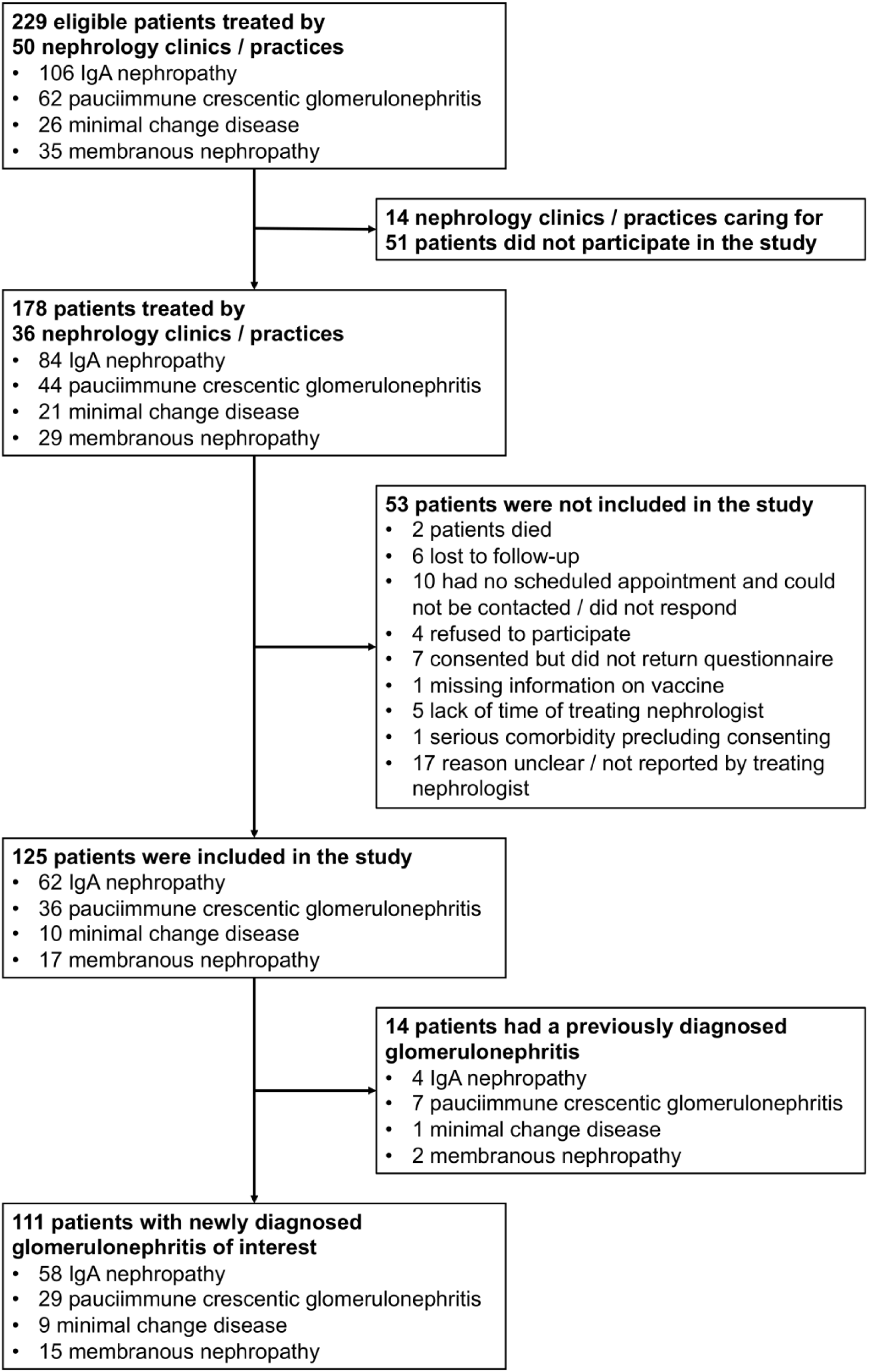
Study flow chart of the second study.

**Table 2.** Clinical characteristics of patients with newly diagnosed glomerulonephritis during the study period.

|  | All Patients | IgAN | PINGN | MCD | MN |
| --- | --- | --- | --- | --- | --- |
| n | 111 | 58 | 29 | 9 | 15 |
| Median age (IQR), y | 55 [43, 72] | 47 [37, 54] | 70 [57, 75] | 70 [61, 72] | 66 [60, 76] |
| Female sex | 42 (38) | 21 (36) | 12 (41) | 5 (56) | 4 (27) |
| **Clinical syndrome** |  |  |  |  |  |
| Nephrotic syndrome | 29 (26) * | 5 (9) * | 0 (0) | 9 (100) | 15 (100) * |
| Nephritic syndrome | 60 (54) * | 30 (52) * | 27 (93) | 0 (0) | 3 (20) * |
| Acute glomerulonephritis | 21 (19) | 7 (12) | 13 (45) | 0 (0) | 1 (7) |
| Chronic glomerulonephritis | 24 (22) | 21 (36) | 1 (3) | 0 (0) | 2 (13) |
| RPGN | 15 (14) | 2 (3) | 13 (45) | 0 (0) | 0 (0) |
| Asymptomatic urinary abnormalities | 25 (23) | 23 (40) | 2 (7) | 0 (0) | 0 (0) |
| Asymptomatic microhematuria | 23 (21) | 21 (36) | 2 (7) | 0 (0) | 0 (0) |
| Asymptomatic proteinuria | 22 (20) | 20 (34) | 2 (7) | 0 (0) | 0 (0) |
| Isolated macrohematuria | 2 (2) | 2 (3) | 0 (0) | 0 (0) | 0 (0) |
| **Laboratory parameters at the time of biopsy** |  |  |  |  |  |
| Creatinine, $\mu\text{mol/L}$, median (IQR) | 128 [85, 190] | 127 [85, 165] | 162 [103, 276] | 130 [104, 263] | 86 [82, 173] |
| eGFR median (IQR) | 48 [29, 72] | 52 [37, 79] | 31 [15, 52] | 47 [15, 66] | 67 [38, 75] |
| Serum albumin g/l median (IQR) | 34 [28, 40] | 39 [36, 43] | 34 [29, 39] | 21 [20, 24] | 25 [22, 28] |
| Proteinuria g/d median (IQR) | 2 [1, 5] | 1 [1, 3] | 1 [0, 3] | 11 [5, 13] | 8 [6, 11] |
| **History of COVID-19** |  |  |  |  |  |
| COVID-19 before biopsy | 8 (7) | 4 (7) | 2 (7) | 1 (11) | 1 (7) |
| COVID-19 before onset of symptoms/signs | 4 (4) | 1 (2) | 2 (7) | 1 (11) | 0 (0) |
| **Vaccination history** |  |  |  |  |  |
| Any vaccine before biopsy | 43 (39) | 20 (34) | 11 (38) | 5 (56) | 7 (47) |
| Any vaccine before onset of signs or symptoms | 22 (20) | 8 (14) | 5 (17) | 4 (44) | 5 (33) |
| Any vaccine within 4 weeks of biopsy | 13 (12) | 9 (16) | 2 (7) | 2 (22) | 0 (0) |
| Any vaccine within 4 weeks of symptom or sign onset | 12 (11) | 6 (10) | 3 (10) | 1 (11) | 2 (13) |

Abbreviations: IgAN, IgA nephropathy; PINGN, pauci-immune necrotizing glomerulonephritis; MCD, minimal change disease; MN, membranous nephropathy; RPGN, rapidly progressive glomerulonephritis.
Values are numbers (percentages) or median (interquartile range).
To convert the creatinine values from $\mu\text{mol/L}$ to $\text{mg/dL}$ , divide by 88.4.
\* 5 patients (2 IgAN and 3 MN) had nephrotic-nephritic syndrome and were counted in both categories.

### Vaccination history of patients with new-onset glomerulonephritis and matched controls

43 (38.7%) of the patients with a newly established histological diagnosis of glomerulonephritis between January and August 2021 had received at least one dose of an mRNA SARS-CoV-2 vaccine prior to kidney biopsy. In 21 of these patients, symptoms or laboratory abnormalities attributable to the glomerulonephritis or extrarenal manifestations of vasculitis had been present before receiving the first vaccine dose. Hence, 22 (19.8%) of the patients with a newly established histological diagnosis of glomerulonephritis had received at least one vaccine dose before onset of symptoms or laboratory abnormalities. The vaccination rates of the general population, matched for age and either date of biopsy or date of onset of symptoms or laboratory abnormalities, were similar (39.5% and 18.4%, respectively). The estimated risk ratios for the development of new-onset biopsy-proven glomerulonephritis and for the development of new symptoms or laboratory abnormalities attributable to glomerulonephritis were 1.03 (95%-confidence interval 0.72 to 1.48, *P*=0.96) and 1.14 (95%CI 0.74 to 1.77, *P*=0.62), respectively, in patients having received at least one dose of SARS-CoV-2 vaccine, compared to unvaccinated persons matched for age and calendar date of biopsy or onset of symptoms or laboratory abnormalities, respectively. Risk ratios for the individual types of glomerulonephritis are shown in **Table 3**.

**Table 3.** Estimated risk ratios for the development of each type of biopsy-proven glomerulonephritis and for the development of new symptoms or laboratory abnormalities attributable to the respective disease.

| **Diagnosis** | **Risk ratio for biopsy proven glomerulonephritis** | **Risk ratio for symptoms or laboratory signs of glomerulonephritis** |
| --- | --- | --- |
| IgAN | 1.14 (95%CI 0.67 to 1.97), $P=0.73$ | 1.14 (95% CI 0.54 to 2.41), $P=0.88$ |
| PINGN | 0.54 (95% CI 0.26 to 1.15), $P=0.15$ | 0.61 (95% CI 0.23 to 1.60), $P=0.42$ |
| MCD | 1.72 (95% CI 0.46 to 6.38), $P=0.63$ | 2.20 (95% CI 0.59 to 8.18), $P=0.41$ |
| MN | 1.17 (95% CI 0.43 to 3.23), $P=0.96$ | 1.64 (95% CI 0.56 to 4.79), $P=0.54$ |
| Any GN | 0.97 (95% CI 0.66 to 1.42), $P=0.95$ | 1.10 (95%CI 0.69 to 1.75, $P=0.79$) |

Abbreviations: IgAN, IgA nephropathy; PINGN, pauci-immune necrotizing glomerulonephritis; MCD, minimal change disease; MN, membranous nephropathy; GN, glomerulonephritis

**Figure 3** shows the percentage of individuals having received any vaccine dose in each four-week time period prior to kidney biopsy or prior to onset of symptoms or laboratory abnormalities, as compared to matched control groups. The analysis by type of glomerulonephritis is shown in **Figure 4**, the analysis by first or second vaccine dose in Supplementary Figure 3.

**Fig 3.**
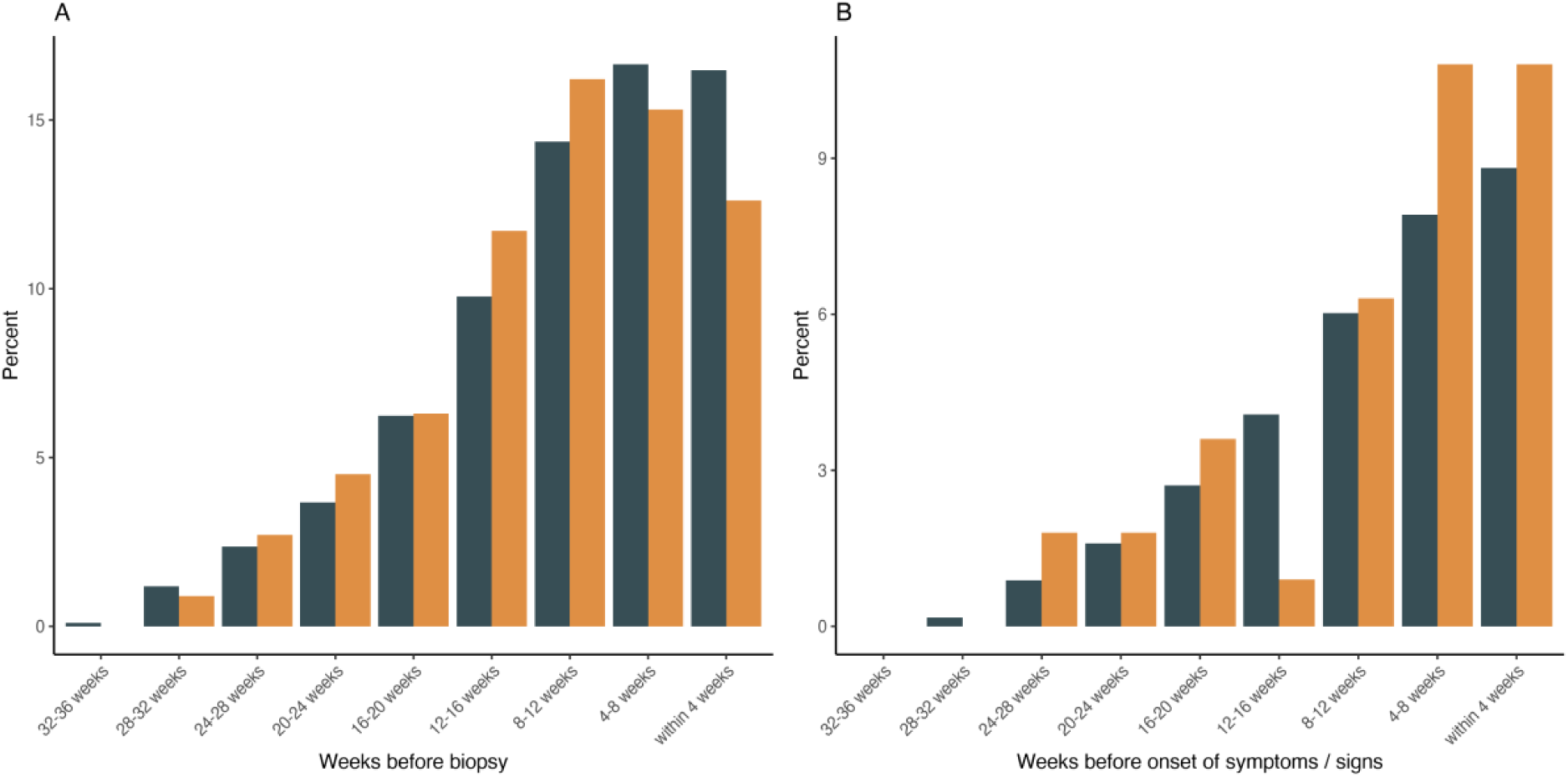
Frequency and timing of vaccination in patients with glomerulonephritis compared to matched controls. Shown in yellow is the percentage of patients with a new diagnosis of IgAN, PINGN, MCD or MN during the study period, who have received any vaccine dose during each four-week interval before renal biopsy (A) or onset of symptoms or signs attributable to the renal disease or an extrarenal manifestation thereof (B). For comparison, the percentage of persons from the general population matched for age and timepoint during the vaccination campaign are shown in dark grey.

**Fig 4.**
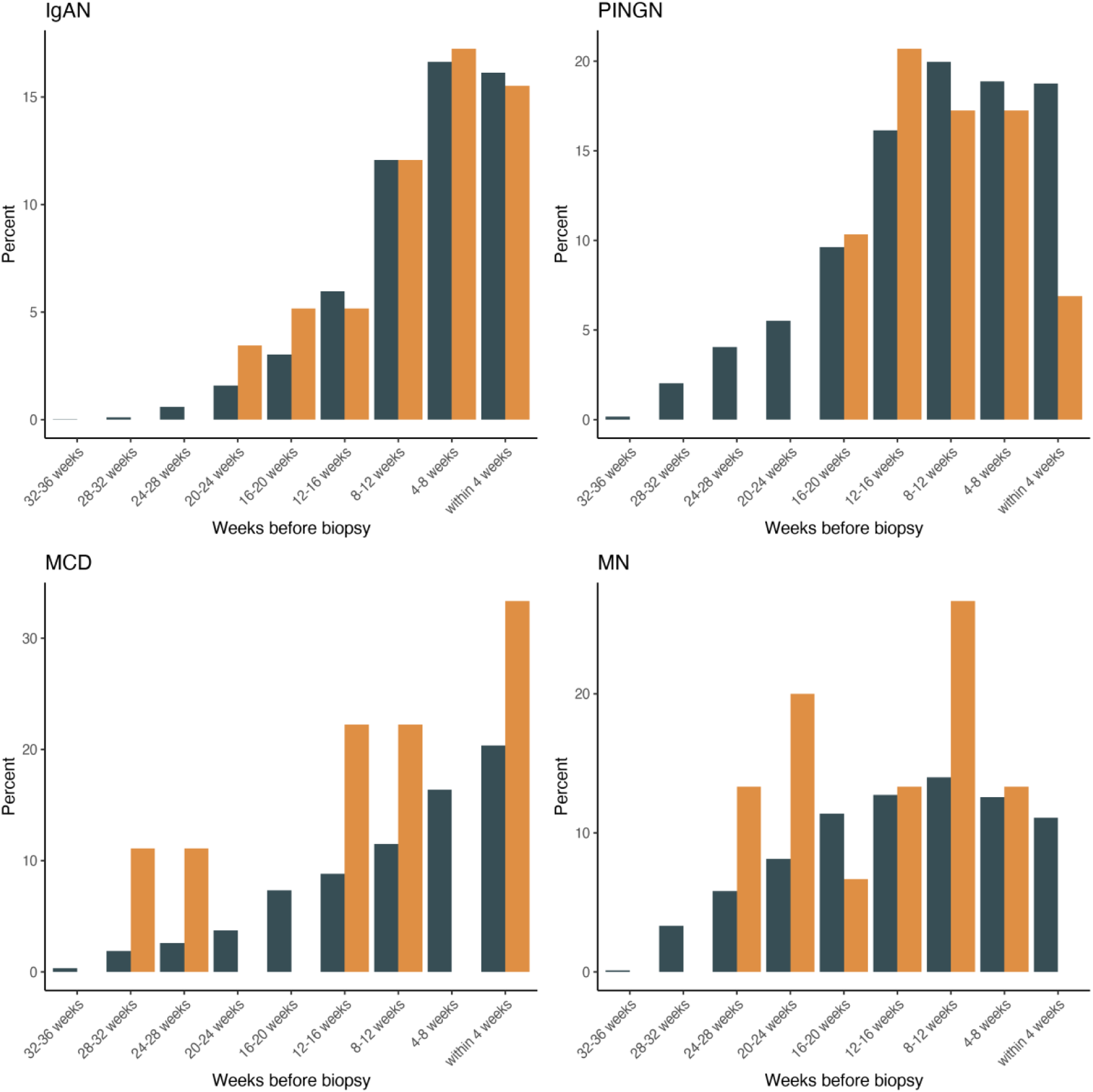
Frequency and timing of vaccination by histological diagnosis in patients with glomerulonephritis compared to matched controls. Shown in yellow is the percentage of patients with a new diagnosis of IgAN (A), PINGN (B), MCD (C) or MN (D) during the study period, who have received any vaccine dose during each four-week interval before renal biopsy. For comparison, the percentage of persons from a control population matched for age and timepoint during the vaccination campaign are shown in grey.

### Clinical characteristics of patients with a diagnosis of glomerulonephritis in temporal association with vaccination

Temporal association of glomerulonephritis with SARS-CoV-2 vaccination has not been uniformly defined in case reports and case series, with the largest case series defining it as onset of symptoms within one month of any vaccine dose.^6^ In 15 patients of our cohort, glomerulonephritis definitely (n=4) or possibly (n=11) manifested within 28 days of a vaccine dose. The clinical characteristics of these patients compared to all other patients with a diagnosis of glomerulonephritis during the study period are shown in **Table 4**. Details on each individual patient are given in Supplementary Table 3. Patients with a new diagnosis of glomerulonephritis manifesting in temporal association with vaccination were older, but otherwise did not differ from patients with a new diagnosis of glomerulonephritis temporally unrelated to SARS-CoV-2 vaccination.

**Table 4.**
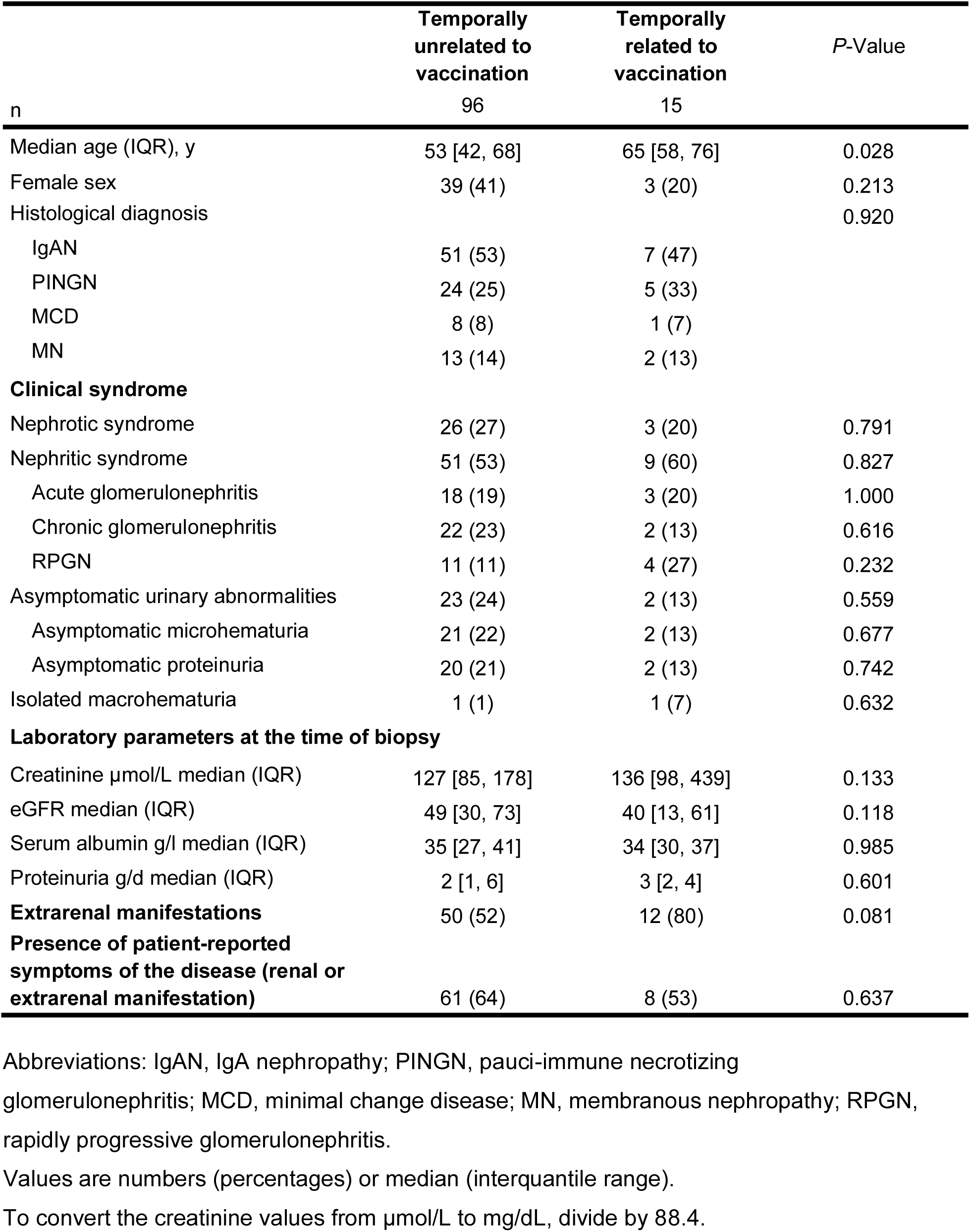
Characteristics of patients with newly diagnosed glomerulonephritis with vs. without temporal association to SARS-CoV-2 vaccination.

|  | Temporally<br>unrelated to<br>vaccination | Temporally<br>related to<br>vaccination | P-Value |
| --- | --- | --- | --- |
| n | 96 | 15 |  |
| Median age (IQR), y | 53 [42, 68] | 65 [58, 76] | 0.028 |
| Female sex | 39 (41) | 3 (20) | 0.213 |
| Histological diagnosis |  |  | 0.920 |
| IgAN | 51 (53) | 7 (47) |  |
| PINGN | 24 (25) | 5 (33) |  |
| MCD | 8 (8) | 1 (7) |  |
| MN | 13 (14) | 2 (13) |  |
| **Clinical syndrome** |  |  |  |
| Nephrotic syndrome | 26 (27) | 3 (20) | 0.791 |
| Nephritic syndrome | 51 (53) | 9 (60) | 0.827 |
| Acute glomerulonephritis | 18 (19) | 3 (20) | 1.000 |
| Chronic glomerulonephritis | 22 (23) | 2 (13) | 0.616 |
| RPGN | 11 (11) | 4 (27) | 0.232 |
| Asymptomatic urinary abnormalities | 23 (24) | 2 (13) | 0.559 |
| Asymptomatic microhematuria | 21 (22) | 2 (13) | 0.677 |
| Asymptomatic proteinuria | 20 (21) | 2 (13) | 0.742 |
| Isolated macrohematuria | 1 (1) | 1 (7) | 0.632 |
| **Laboratory parameters at the time of biopsy** |  |  |  |
| Creatinine $\mu\text{mol/L}$ median (IQR) | 127 [85, 178] | 136 [98, 439] | 0.133 |
| eGFR median (IQR) | 49 [30, 73] | 40 [13, 61] | 0.118 |
| Serum albumin g/l median (IQR) | 35 [27, 41] | 34 [30, 37] | 0.985 |
| Proteinuria g/d median (IQR) | 2 [1, 6] | 3 [2, 4] | 0.601 |
| **Extrarenal manifestations** | 50 (52) | 12 (80) | 0.081 |
| **Presence of patient-reported symptoms of the disease (renal or extrarenal manifestation)** | 61 (64) | 8 (53) | 0.637 |

Abbreviations: IgAN, IgA nephropathy; PINGN, pauci-immune necrotizing glomerulonephritis; MCD, minimal change disease; MN, membranous nephropathy; RPGN, rapidly progressive glomerulonephritis.
Values are numbers (percentages) or median (interquartile range).
To convert the creatinine values from $\mu\text{mol/L}$ to $\text{mg/dL}$ , divide by 88.4.

### Sensitivity analysis

The sensitivity analysis excluding all MN cases showed equivalent results (Supplementary Tables 1, 4 and 5, Supplementary Figures 4 and 5).

## Discussion

Postmarketing surveillance is an important means to detect rare but relevant side-effects that may have escaped detection in registration studies. However, while serving an important hypothesis-generating function, case reports and series are subject to post hoc fallacy and publication bias. In this study, which included incidence data of glomerulonephritis for 7.1 million individuals, 69% of whom had received at least one vaccine dose during the study period, we did not find evidence that mRNA-based vaccines against SARS-CoV-2 increase the risk for new-onset glomerulonephritis. On the epidemiological level, incidence of four common types of glomerulonephritis did not exceed the expected rate during any month of the SARS-CoV-2 vaccination campaign in Switzerland. Patients with new-onset glomerulonephritis did not differ from age- and calendar-time-matched controls with respect to their vaccination history, and characteristics of patients with glomerulonephritis temporally related to vaccination did not differ from those without temporal association to vaccination, except for higher age, probably due to preferential vaccination of older individuals.

Glomerulonephritis is a rare disease and nationwide incidence rates have not been previously published for Switzerland. We found baseline incidence rates similar to those reported for other European countries.^17-19^ An interesting finding was the significantly reduced incidence of glomerulonephritis during public lockdown measures paralleling the first infection wave. Notably, this reduction was attributable to IgAN, the most frequent of the four glomerulonephritides, which often manifests with isolated, asymptomatic urinary abnormalities not necessitating urgent evaluation and more likely to be missed with reduced use of routine medical evaluations during the first pandemic phase. Importantly, this finding demonstrates the feasibility of our study to detect true changes of incidence.

Patients with a new diagnosis of histology-proven glomerulonephritis had similar vaccination histories compared to the matched population-based cohort, and the estimated risk for the development of glomerulonephritis was equal in vaccinated vs. unvaccinated persons. In the time-based analysis, the proportion of patients having received a vaccine dose was virtually identical for every four-week period before histological diagnosis of a glomerulonephritis, compared to the matched control cohort. A trend towards less vaccines given to patients within four weeks before histological diagnosis of glomerulonephritis is likely attributable to the fact that patients were less likely to schedule a vaccine appointment shortly before a scheduled kidney biopsy or when suffering symptoms of glomerulonephritis or vasculitis. In the two four-week-periods before symptom onset or first documented laboratory abnormalities, the percentages of patients having received a vaccine dose were nominally slightly higher compared to matched controls. However, this analysis has to be interpreted with caution, because the number of patients was relatively low and some patient-reported symptoms are unspecific and may have incorrectly been attributed to glomerulonephritis.

Our findings were generally consistent across all four types of glomerulonephritis, with the possible exception of MCD. Increased numbers of newly diagnosed MCD were recently reported for 2021 in a Dutch observational cohort.^20^ However, that cohort-based study could not calculate true population-based incidence, and only 5 of 11 cases had received any SARS-CoV-2 vaccine before presentation, which makes a true association with vaccination questionable. In our study, the incidence of MCD during the vaccination campaign did not cross the upper boundary of the 95% credible interval for the predicted incidence, and the risk ratio for the development of MCD in vaccinated vs. unvaccinated persons was not significantly different from one. However, both analyses showed a trend towards an increased risk, and 95% credible / confidence intervals were wide due to the overall low incidence of MCD. Thus, we cannot exclude an effect of SARS-CoV-2 mRNA vaccines on the development of MCD, but the absolute risk would be very small.

We identified 15 patients with new-onset glomerulonephritis in possible temporal association to SARS-CoV-2 vaccination. While the limited number of cases precluded a detailed analysis, we did not find a distinct clinical manifestation in these patients. Macrohematuria has been frequently reported as clinical manifestation in patients with new onset or relapsing glomerulonephritis temporally associated with SARS-CoV-2 vaccination.^21^ Episodes of gross hematuria, often in association with upper respiratory tract infection, are a known manifestation of IgA nephropathy^22^ and could theoretically be triggered by the systemic inflammatory response following mRNA-based SARS-CoV-2 vaccination. However, we did not find an increased rate of patient-reported gross hematuria in patients with newly diagnosed IgAN in temporal association to vaccination compared to the rest of the cohort. Furthermore, among a previously reported cohort of 88 patients with known IgA nephropathy who received at least one dose of mRNA based SARS-CoV-2 vaccine, none had developed gross hematuria.^23^

Strengths of this study are the inclusion of all biopsy-proven glomerulonephritis cases diagnosed in Switzerland, which allowed to calculate true incidence rates, and the use of an unambiguous primary outcome (biopsy-proven glomerulonephritis) that limited the potential for bias. By linking the second study to the epidemiological analysis, we were able to specifically approach potential participants and achieved a considerable inclusion rate (>50% of all Swiss patients with histologically confirmed glomerulonephritis during the study period). The availability of detailed information on vaccinations administered to the general Swiss population allowed precise matching to patients with regards to age and timepoint during the vaccination campaign. Since only mRNA-based vaccines were administered in Switzerland during the study period, our study specifically addresses this type of vaccines.

This study has several limitations. First, due to the limited population size of Switzerland and the low incidence of glomerulonephritis, we cannot exclude a small effect of vaccinations, in particularly for MCD, as discussed above. Second, due to the decentralized health care system in Switzerland, patients were cared for by a variety of hospital-based and private practice nephrologists. Patient evaluation and care were thus not standardized and the study relied on patient- and physician-reported data. Third, due to the retrospective design of the study, patient-reported symptoms were subject to potential recall bias and onset of symptom dates were not precise in some patient questionnaires. Therefore, we chose biopsy proven glomerulonephritis as the primary outcome. Fourth, we were not able to include all patients with biopsy-proven glomerulonephritis in the second study and cannot exclude selection bias. However, the major reason for non-inclusion of patients was non-participation of their treating nephrology divisions or practices due to time constraints, which should not introduce bias. Finally, our results are limited to new-onset glomerulonephritis and cannot answer the question, whether SARS-CoV-2 vaccination could trigger relapses in patients with previously diagnosed glomerulonephritis, because relapses are usually diagnosed clinically without repeat biopsy.

In conclusion, combining two complementary approaches, we did not find an association between mRNA-based vaccination against SARS-CoV-2 and an increased incidence of the four common glomerulonephritis types IgAN, PINGN, MCD and MN. Most cases of new onset glomerulonephritis manifesting shortly after vaccination against SARS-CoV-2 are likely attributable to temporal coincidence.

## Data Availability

Data will be made available on reasonable request to the corresponding author, after approval by the local ethics committee.

## Author contributors

A. D. Kistler, E. Locher and M. Diebold had full access to all the data in the study and take responsibility for the integrity of the data and the accuracy of the data analysis. Kistler developed the concept and designed the study and drafted the manuscript. Diebold and Kistler did the statistical analysis. All authors contributed to the acquisition, analysis, and interpretation of data; and the critical revision of the manuscript for important intellectual content. The corresponding author attests that all listed authors meet authorship criteria and that no others meeting the criteria have been omitted.

## Additional contributions

We thank Marcel Zwahlen and Julien Riou, both from the Institute of Social and Preventive Medicine, University of Bern, Bern, Switzerland, for epidemiological and statistical support, in particular with the Bayesian regression model.Menno Prujim, Service of Nephrology and Hypertension, Department of Medicine, Lausanne University Hospital and University of Lausanne, Switzerland, helped create the local biopsy database at the University of Lausanne and supervised G. Nanchen.

The following nephrologists from the indicated nephrology divisions and practices – in addition to those listed among the authors – have recruited patients for the second study and provided their clinical data:

Bürgerspital Solothurn: S. Zschiedrich; CHUV: M. Stevanin; Dialyse Riviera (Vevey): T. Gauthier, R. Chazot; Ente Ospedaliero Cantonale (Lugano): G. Bedino; Gesundheitszentrum Fricktal: T. Öttl; Herz-und Nierenzentrum Aare (Solothurn): S. Farese; Hirslanden Klinik St. Anna (Luzern): A. Jehle; Hôpital du Jura (Delémont): P. Wilson; Hôpital Riviera Chablais (Rennaz): A. Rossier; Hôpitaux Universitaires de Genève: A. Faivre; Hôpital du Valais (Sion) : G. Guzzo ; Kantonsspital Baden: C. Gussone; Kantonsspital Baselland: F. Burkhalter, Y. Holzmann, C. Jäger, S. Kalbermatter; Kantonsspital Frauenfeld: D. Daiss, A. Keil, S. Flury; Kantonsspital Graubünden: A. Georgalis; Kantonsspital Olten: C. Lenherr; Kantonsspital Uri: D. Bruhin; Kantonsspital Winterthur: S. J. Rippin Wagner, L. Nigg Calanca; Luzerner Kantonsspital: A. Duss, A. Rali, S. Maloney, M. Neher; Nieren-und Dialysezentrum Männedorf: P. Rhyn; Nierenzentrum Rheintal (Altstätten): R. Eisel, C. Jäger; Quavitae Rive Gauche (Genève) : V. Jotterand Drepper; Regionalspital Emmental (Burgdorf): I. Bergmann; Spital Lachen: R. Schorn; Spital Thun: B. Landtwing Lüscher; Spitalverbund Appenzell Auserrhoden: I. Koneth, T. Staub; Spitalzentrum Biel: N. Drivakos, A. Kruse; Spitalzentrum Oberwallis (Visp): D. Brodmann, C. Brun; Spital Zollikerberg: B. Bergamin, M. Pechula Thut; Stadtspital Waid und Triemli (Zürich): A. Helmuth, A. Schleich, N. Weber, J. Meier; Universitätsspital Basel: M. Dickenmann, P. Hirt-Minkowski.

