## Supplementary Material for "Incidence of Glomerulonephritis after SARS-CoV-2 mRNA Vaccination"

#### Supplementary methods

**Data collection for the second study**

**List of the used packages in R**

#### Supplementary Tables

**Supplementary Table 1.** Weekly incidence of histologically diagnosed glomerulonephritis per 1'000'000 during the entire study period and during the peak of the vaccination campaign, compared to the corresponding time of the year in the baseline period.

**Supplementary Table 2.** Clinical details and indications for repeat biopsy of patients with previously known glomerulonephritis.

**Supplementary Table 3.** Clinical details on glomerulonephritis cases temporally related to SARS-CoV-2 vaccination

**Supplementary Table 4.** Observed monthly incidence of glomerulonephritis (excluding membranous nephropathy) during the vaccination campaign compared to the baseline period (2015 – 2019) and the expected incidence, relative risk ratio and number of vaccine doses administered in the corresponding month.

**Supplementary Table 5.** Estimated risk ratio for the development of biopsy-proven glomerulonephritis (excluding membranous nephropathy) and for the development of new symptoms or laboratory abnormalities excluding patients with membranous nephropathy.

#### Supplementary Figures

**Supplementary Figure 1.** Monthly incidence of IgAN, PINGN, MCD and MN from January 2015 – August 2021.

**Supplementary Figure 2.** Observed and expected incidence of glomerulonephritis during 2020, the first pandemic year.

**Supplementary Figure 3.** Frequency and timing of vaccination by dose number in patients with glomerulonephritis compared to matched controls.

**Supplementary Figure 4.** Observed and expected incidence of glomerulonephritis during the vaccination campaign, excluding membranous nephropathy.

**Supplementary Figure 5.** Frequency and timing of vaccination in patients with glomerulonephritis (excluding membranous nephropathy) compared to matched controls.

### Supplementary Methods

#### Data collection for the second study

The following informations were collected in the case report form: biopsy date, patient age, gender; histological diagnosis with free text for details, comments and results of serology; newly established diagnosis vs. repeat biopsy; clinical syndrome (nephrotic syndrome, nephritic syndrome further specified as acute glomerulonephritis, chronic glomerulonephritis or rapidly progressive glomerulonephritis, asymptomatic proteinuria, asymptomatic microhematuria, macrohematuria, other); serum creatinine, serum albumin, proteinuria and urinary sediment (both at the time of renal biopsy and worst value within +/- one month of biopsy); date of the first documentation of hematuria, proteinuria >1g/day, proteinuria >3.5g/day, hypoalbuminemia, and worsening renal function; extrarenal manifestations (arthralgia / arthritis, rash, ENT-manifestations, pulmonary manifestations, GIT-manifestations, neurological manifestations, other, all to be specified with free text); comorbidities (diabetes mellitus with year of onset, hypertension with year of onset, coronary artery disease, chronic obstructive pulmonary disease, other).

Proteinuria was provided by the treating nephrologists either in in gram per day from a 24h urine collection or as the protein-to-creatinine ratio in g/mmol from a spot urine sample. The latter was multiplied with ten to obtain an estimate of 24h proteinuria.

The following informations were collected in the patient questionnaire: patient age and gender; symptoms of the renal disease (none; bloody / brown urine, foamy urine, edema, arthralgia / arthritis, rash, cough, shortness of breath, abdominal pain, bloody diarrhea, ocular inflammation, nasal congestion / bloody nasal discharge, focal paralysis / sensory loss, other; each with date of first occurrence); information on SARS-CoV-2 infection (yes / no, date of symptom onset and date of positive testing), history of vaccine against SARS-CoV-2 (type of vaccine and date of doses received).

All data were reviewed by the investigators. The classification of the clinical syndrome was primarily based on the treating nephrologists' judgement on the CRF as well as on the laboratory data on the CRF (e.g. where "asymptomatic microhematuria" was ticked in addition to "nephritic syndrome", the former was not accounted for since microhematuria is part of the nephritic syndrome). The date of onset of symptoms or first detection of laboratory anomalies was independently adjudicated by two investigators blinded to vaccination history of the patients. We used the following rules: first detection of laboratory anomalies was defined by the date of first occurrence (according to the CRF) of hematuria, proteinuria >1g/day, or worsening renal function (whatever occurred first), if the histological diagnosis was IgAN or PINGN; and by the date of first occurrence of proteinuria >1g/day, or hypoalbuminemia (whatever occurred first), if the histological diagnosis was MN or MCD, unless hypoalbuminemia was clearly attributable to a comorbidity (e.g. liver cirrhosis). Onset of symptoms was defined by the first occurrence (according to the patient questionnaire) of renal or extrarenal symptoms attributable to the glomerular disease. With respect to renal symptoms, if the histological diagnosis was PINGN, patient-reported macrohematuria and edema (ankle / leg swelling) was attributed to the glomerular disease; if the histological diagnosis was IgAN, patient-reported macrohematuria was attributed to the glomerular disease, and edema only if nephrotic syndrome was present; if the histological diagnosis was MCD or MN, edema and foamy urine were attributed to the glomerular disease. If any of the above symptoms were attributable to a comorbidity and clearly predated the onset of other symptoms / signs, they were not attributed to the renal disease. Extrarenal symptoms reported by the patients (and their date of onset) were only attributed to the glomerular disease in PINGN as part of systemic AAV and in IgAN as part of IgA vasculitis. Because many of these symptoms are unspecific, patient-reported symptoms were only attributed to the glomerular disease, if a corresponding extrarenal manifestation of the disease was reported on the CRF by the treating nephrologists. In cases where patients remembered only the month of symptom onset, the date of onset was set to the 15$^{th}$ day of that month.

#### List of the used packages in R

- readxl
- finalfit
- dplyr
- ggplot2
- tidyr
- tableone
- tibble
- ggpubr
- readr
- lubridate
- slider
- timetk
- rstanarm
- epitools

**Supplementary Table 1. Weekly incidence of histologically diagnosed glomerulonephritis per 1'000'000 during the entire study period and during the peak of the vaccination campaign, compared to the corresponding time of the year in the baseline period.**

| | **Weekly incidence** | | | | | | |
|---|---|---|---|---|---|---|---|
| | Entire year | January - August | | | May - August | | |
| **Histological diagnosis** | 2015 - 2019 | 2015 - 2019 | 2021 | p-Value | 2015 - 2019 | 2021 | p-Value |
| IgA nephropathy | 0.48 ± 0.26 | 0.49 ± 0.26 | 0.48 ±0.21 | 0.949 | 0.50 ± 0.30 | 0.46 ± 0.26 | 0.569 |
| Pauciimmune necrotizing glomerulonephritis | 0.29 ± 0.17 | 0.29 ± 0.17 | 0.28 ± 0.12 | 0.611 | 0.26 ± 0.14 | 0.27 ± 0.14 | 0.707 |
| Minimal change disease | 0.20 ± 0.10 | 0.20 ± 0.09 | 0.22 ± 0.12 | 0.634 | 0.21 ± 0.10 | 0.21 ± 0.11 | 0.938 |
| Membranous nephropathy | 0.25 ± 0.14 | 0.25 ± 0.13 | 0.21 ± 0.09 | 0.046 | 0.24 ± 0.11 | 0.15 ± 0.04 | <0.001 |
| Any glomerulonephritis of interest | 0.96 ± 0.42 | 0.95 ± 0.42 | 0.92 ± 0.36 | 0.636 | 0.88 ± 0.32 | 0.84 ± 0.35 | 0.669 |
| Any glomerulonephritis excluding MN | 0.78 ± 0.36 | 0.78 ± 0.38 | 0.80 ± 0.32 | 0.688 | 0.73 ± 0.38 | 0.74 ± 0.36 | 0.871 |

#### Supplementary Table 2. Clinical details and indications for repeat biopsy of patients with previously known glomerulonephritis

| Pat Nr | Age decade (y) | Sex | Histological diagnosis | Clinical manifestations | | Patient-reported gross hematuria | Laboratory parameters | | | | History of COVID-19 | Vaccination history | Biopsy date | Vaccine before biopsy |
|---|---|---|---|---|---|---|---|---|---|---|---|---|---|---|
| | | | | renal | extrarenal | | crea | eGFR | Serum albumin | U Prot | | | | |
| 1 | 70-79 | m | PINGN | cGN | unspecific | yes | 335 | 15 | n/a | 1.9 | no | M, 03/21, 03/21 | 08/21 | yes |
| 2 | 60-69 | m | PINGN | cGN | no | no | 440 | 12 | 23 | 16.3 | no | M, 02/21, 03/21 | 05/21 | yes |
| 3 | 50-59 | m | PINGN | aMH&P | no | no | 91 | 87 | 38 | 2.0 | no | P, 04/21, 05/21 | 06/21 | yes |
| 4 | 40-49 | f | MN | NS | no | no | 51 | 111 | 31 | 5.0 | no | P, 01/21, 02/21 | 01/21 | yes |
| 5 | 40-49 | f | IgAN | IgAN | skin | no | 56 | 109 | n/a | 1.4 | 12/21 | P, 03/21, 04/21 | 05/21 | yes |
| 6 | 50-59 | m | IgAN | cGN | skin | no | 148 | 47 | n/a | 4.0 | 11/20 | M, 06/21 | 06/21 | yes |
| 7 | 40-49 | m | PINGN | cGN | no | no | 127 | 60 | n/a | 0.5 | 10/21 | P, 03/21, 04/21 | 06/21 | yes |
| 8 | 30-39 | m | PINGN | cGN | no | no | 179 | 42 | 41 | 1.7 | no | M, 06/21, 07/21 | 04/21 | no |
| 9 | 30-39 | m | MCD | aP/NS | no | no | 74 | 119 | 29 | 5.2 | no | no | 06/21 | no |
| 10 | 80-89 | m | IgAN | cGN | no | no | 252 | 22 | 29 | 0.7 | no | M, 03/21, 04/21 | 03/21 | yes |
| 11 | 30-39 | f | IgAN | cGN | no | no | 197 | 29 | 38 | 4.4 | 12/20 | no | 03/21 | no |
| 12 | 50-59 | m | PINGN | NS | ENT | no | 240 | 27 | 42 | 3.3 | no | P, 03/21, 04/21 | 03/21 | yes |
| 13 | 70-79 | m | PINGN | cGN | ENT | no | 81 | 89 | 37 | 0.4 | no | M, 01/21, 02/21 | 01/21 | yes |
| 14 | 60-69 | f | MN | aP | no | no | 64 | 93 | 41 | 0.8 | no | P, 03/21, 04/21 | 03/21 | no |

Abbreviations: crea, serum creatinine (in μmol/l); eGFR, estimated glomerular filtration rate according to the CKD-EPI 2009 formula (in ml/min/1.73m$^2$); U Prot, urinary protein excretion in g per day, measured from a 24h urinary collection or estimated from the protein-creatinine-ratio; M, Moderna mRNA-1273 vaccine; P, Pfizer-BioNTech BNT162b2 vaccine; n/a, not available; IgAN, IgA nephropathy; PINGN, pauci-immune necrotizing glomerulonephritis; MCD, minimal change disease; MN, membranous nephropathy; RPGN, rapidly progressive glomerulonephritis; NS, nephrotic syndrome; aGN / cGN, acute / cronic glomerulonephritis; aHM&P, asymptomatic microscopic hematuria and proteinuria; ENT, ears nose throat. Dates are given as month/year. Dates under History of COVID-19 refer to date of symptom onset.

Additional details on case presentations:

**Patient 1**: Diagnosis of MPO-ANCA Vasculitis in September 2009. Renal flare with nephritic syndrome in August 2021.

**Patient 2**: First diagnosis of ANCA-vasculitis in 2011. Patient suffered acute kidney injury requiring dialysis. Renal biopsy was performed to exclude alternative cause of AKI and showed active (crescents) and chronic lesions.

**Patient 3**: Patient with new-onset proteinuria and hematuria. First diagnosis 2015. In renal biopsy active and chronic lesions.

**Patient 4**: First biopsy in 1996. In January 2021 new increase in proteinuria.

**Patient 5**: In 2014 diagnosis of IgA Vasculitis by skin and kidney biopsy. In June 2020 new diagnosis of systemic lupus erythematodes. Kidney biopsy was performed to check for concomitant Lupus Nephritis. No signs of Lupus Nephritis were seen in kidney biopsy.

**Patient 6**: Diagnosis of IgA Vasculitis in 2008. New worsening proteinuria.

**Patient 7**: Evaluation for slowly worsening renal function with proteinuria and microhematuria in the setting of known ANCA-associated vasculitis, currently without symptoms. A first kidney biopsy was performed before 2015 abroad and the original report was not available, therefore, a repeat biopsy was performed.

**Patient 8**: Known diagnosis since 2000. Indication for kidney biopsy was worsening of renal function.

**Patient 9**: Features consistent with MCD, but hereditary podocytopathy considered as an alternative diagnosis. Indication for kidney biopsy was worsening of proteinuria with incomplete nephrotic syndrome (nephrotic-range proteinuria, hypoalbuminemia but no edema).

**Patient 10**: Known IgAN with chronic kidney disease. Indication for repeat biopsy was not reported by the treating physician.

**Patient 11**: Known IgAN since 2014. Indication for repeat biopsy was worsening renal function.

**Patient 12**: Previously-known MPO-ANCA vasculitis since July 2020. Did not tolerate cyclophosphamide. Repeat biopsy performed because of persistent / progressive disease under treatment with rituximab.

**Patient 13**: Patient with known PINGN since 2006. New onset of hematuria.

**Patient 14**: Patient with a diagnosis of membranous nephropathy since 2009. Known history of ANCA-associated vasculitis. Worsening proteinuria and increase in ANCA-titers. Biopsy showed no signs of PINGN.

**Supplementary Table 3. Clinical details on glomerulonephritis cases temporally related to SARS-CoV-2 vaccination**

| Pat Nr | Age decade (y) | Sex | Histological diagnosis | Clinical manifestations | | Patient-reported gross hematuria | Laboratory parameters | | | | History of COVID-19 | Vaccination history | Symptom onset | Abnormal lab values since | Biopsy date |
|---|---|---|---|---|---|---|---|---|---|---|---|---|---|---|---|
| | | | | renal | extrarenal | | crea | eGFR | Serum albumin | U Prot | | | | | |
| **Temporally related cases** | | | | | | | | | | | | | | | |
| 1 | 80-89 | m | PINGN | RPGN | no / unspecific | no | 614 | 7 | n/a | 17.6 | no | P,02/21,+28 | none | d+34 | d+40 |
| 2 | 60-69 | m | MCD | NS | no | no | 428 | 12 | 28 | 5.4 | 04/21 | M,04/21,+22 | d+8 | d+12 | d+19 |
| 3 | 30-39 | f | IgAN | iMH | no | yes | 45 | 123 | 45 | 2.0 | no | P,06/21,+28 | d+30 | d+30 | d+35 |
| 4 | 50-59 | f | IgAN | aGN | no | yes | 131 | 40 | 28 | 4.1 | no | M,06/21,+28 | d+44 | d+44 | d+48 |
| **Cases with possible temporal relation** | | | | | | | | | | | | | | | |
| 5 | 70-79 | m | MN | NS | no | no | 85 | 75 | 27 | 4.3 | no | P,01/21,+21 | d+31 | d+71 | d+101 |
| 6 | 80-89 | f | PINGN | aGN | pulmonary | no | 98 | 45 | 34 | 4.0 | no | M,03/21,+28 | unclear | d+36 | d+40 |
| 7 | 50-59 | m | IgAN | cGN | no | no | 176 | 36 | n/a | 7.1 | no | P,05/21,+28 | none | d+27 | d+37 |
| 8 | 70-79 | m | IgAN | cGN | no | no | 274 | 19 | 38 | 2.9 | no | M,01/21,+29 | none | d+42 | d+152 |
| 9 | 60-69 | m | IgAN | aMH&P | skin | no | 128 | 52 | 33 | 6.8 | no | P,04/21,+64 | 04/21 | d+49 | d+78 |
| 10 | 50-59 | m | PINGN | RPGN | multiple | no | 136 | 50 | 29 | 0.4 | no | M,04/21,+28 | unclear | d+55 | d+75 |
| 11 | 20-29 | m | IgAN | aGN | no | no | 450 | 15 | 33 | 2.7 | no | M,06/21,+34 | unclear | d+8 | d+15 |
| 12 | 70-79 | m | PINGN | RPGN | probably no | no | 756 | 6 | 36 | 3.4 | no | P,04/21,+33 | unclear | d+89 | d+92 |
| 13 | 60-69 | m | IgAN | aMH&P | no | no | 97 | 70 | 45 | 0.2 | no | P,04/21,+30 | none | d+32 | d+77 |
| 14 | 70-79 | m | PINGN | RPGN | no | no | 1180 | 3 | 34 | 1.7 | 02/21 | M,06/21 | none | d+27 | d+29 |
| 15 | 80-89 | m | MN | NS | no | no | 86 | 72 | 18 | 7.9 | no | P,03/21,+32 | 05/21 | d+118 | d+144 |

Abbreviations: crea, serum creatinine (in μmol/l); eGFR, estimated glomerular filtration rate according to the CKD-EPI 2009 formula (in ml/min/1.73m$^2$); U Prot, urinary protein excretion in g per day, measured from a 24h urinary collection or estimated from the protein-creatinine-ratio; M, Moderna mRNA-1273 vaccine; P, Pfizer-BioNTech BNT162b2 vaccine; n/a, not available; IgAN, IgA nephropathy; PINGN, pauci-immune necrotizing glomerulonephritis; MCD, minimal change disease; MN, membranous nephropathy; RPGN, rapidly progressive glomerulonephritis; NS, nephrotic syndrome; aGN / cGN, acute / cronic glomerulonephritis; aHM&P, asymptomatic microscopic hematuria and proteinuria; iMH, isolated macrohematuria. Dates are given as year/month for the first vaccine dose and as days after the first vaccine dose for the second dose and for time points of symptom onset (where exactly known) / first detected laboratory anomalies, and biopsy. Dates under history of COVID-19 refer to month/year of symptom onset.

Additional details on case presentations:

**Patient 1:** Presentation with malaise, nausea and myalgia (date of onset unclear), laboratory diagnosis of renal failure, proteinuria, hematuria and hypoalbuminemia in 03/21, histological diagnosis of PR3-positive ANCA-associated vasculitis.

**Patient 2:** First vaccine dose on in 04/21, COVID-19 with onset of symptoms 7 days after first vaccine dose and positive test 5 days later. The patient first noted edema 8 days after the first vaccine dose, worsening renal function (progressing to acute kidney injury stage 3) was first detected on 10 days, proteinuria and hypoalbuminemia 12 days after the first vaccine dose.

**Patient 3:** Developed fever and arthralgia the day after the second vaccine dose and presented with gross hematuria the day after. Urinalysis revealed hematuria and proteinuria, renal function was normal. Biopsy revealed IgAN with some degree of interstitial fibrosis and tubular atrophy, suggesting preexisting IgAN.

**Patient 4:** Presentation with cough in 07/21, which was due to bilateral atypical pneumonia caused parainfluenza virus type 3. 6 days later, the patient had gross hematuria and laboratory analysis revealed worsening renal function.

**Patient 5:** Onset of edema (swollen ankles) was reported by the patient in 02/21 (does not remember exact date), ca. one month after the first COVID vaccine. Proteinuria and hypoalbuminemia were first documented ca. 1 month later. Serology for PLA2R and THSD7A was negative.

**Patient 6:** Presentation with systemic inflammation. Hematuria first detected in urinary sediment 36 days after the first and 8 days after the second vaccine dose, rapidly worsening renal function after the kidney biopsy, which revealed PINGN but the patient was serologically ANCA negative. CT revealed some nodular infiltrates, probably reflecting pulmonary manifestations of vasculitis. The patient reported cough already since 3 months before the first vaccine, which clearly predated vaccination but is not attributable to vasculitis with certainty.

**Patient 7:** Patient with rheumatoid arthritis since 2007 and methotrexate induced pneumonitis in 2018. Hematuria and worsening renal function were detected in 06/21, i.e. 27 days after the first vaccine dose, but GFR was reduced since December 2016. The syndrome was clinically classified by the treating physician as chronic glomerulonephritis and glomerulonephritis may have preexisted before vaccination.

**Patient 8:** Worsening renal function was first reported 42 days after the first and 11 days after the second vaccine dose, proteinuria and hematuria three months later. The patient was asymptomatic. Biopsy revealed IgAN with signs of chronicity (Oxford classification M1 E0 S1 T1). Thus, worsening renal function was noted within 28 days of the second vaccine dose, but IgAN probably preexisted

**Patient 9:** Presentation with rash / leukocytoclastic vasculitis in 04/21 (unclear if before or shortly after the first vaccine dose), hematuria and low grade proteinuria were first noted two months later in urinalysis and prompted renal biopsy.

**Patient 10:** Suffered from fever and arthralgia the day after the second vaccine dose. After some improvement, unspecific symptoms (malaise), followed by arthralgia and myalgia, cough, dyspnea and nasal congestion with bloody discharge developed. Hematuria was first detected by the general practitioner 55 days after the first and 27 days after the second vaccine dose, renal function started worsening two weeks and proteinuria was first detected three weeks later, PR-3 positivity was found in serology. Pulmonary manifestations were confirmed by CT and mild alveolar hemorrhage by bronchoscopy. The patient reported having suffered from mild cough already three months before receiving the first vaccine dose, which might have represented an early manifestation of vasculitis.

**Patient 11:** Presentation with hypertensive crisis. Worsening renal function, hematuria and proteinuria were detected 8 days after the first vaccine dose. The patient had, however, suffered from headache and blurred vision already one month earlier. Biopsy revealed thrombotic microangiopathy in addition to IgAN. Headache and blurred vision, which preceded the vaccine, may have represented already symptoms of the disease, therefore the timepoint of symptom onset is not entirely clear.

**Patient 12:** Worsening renal function, hematuria and proteinuria were noted in 07/21, 89 days after the first and 56 days after the second vaccine dose. The patient reported shortness of breath since fall 2020 and loss of appetite since 06/21, ca. 4 weeks after the second vaccine dose. While the latter is probably attributable to uremia, it is unclear if dyspnea represented an extrarenal manifestation of vasculitis, but rather unlikely since the treating physician did not report pulmonary manifestations.

**Patient 13:** Indication for the biopsy was hematuria and low-grade proteinuria noted on urinary analysis two days after the second vaccine dose. The patient was asymptomatic.

**Patient 14:** The patient had COVID-19 in January 2021 and received one dose of mRNA-1273 vaccine in 06/21. Worsening renal function and hematuria and proteinuria on urinalysis were noted on 27 days later. The patient reported no symptoms.

**Patient 15:** The patient reported edema starting in 05/21 (exact date not remembered), hence possibly within 28 days of the second vaccine dose. Cough and dyspnea were present since 12/20 (probably not related to nephrotic syndrome, but unclear). In 03/21 he suffered from a leg ulcer (ulcus hypertonicus Martorell). Proteinuria was first documented 118 days after the first and 86 days after the second vaccine dose, hypoalbuminemia one day later.

**Supplementary Table 4. Observed monthly incidence of glomerulonephritis (excluding membranous nephropathy) during the vaccination campaign compared to the baseline period (2015 – 2019) and the expected incidence, relative risk ratio and number of vaccine doses administered in the corresponding month.**

| | **Jan** | **Feb** | **Mar** | **Apr** | **May** | **Jun** | **Jul** | **Aug** |
|---|---|---|---|---|---|---|---|---|
| Incidence 2021 | 3.09 | 2.53 | 4.22 | 4.08 | 4.08 | 3.23 | 3.09 | 2.95 |
| Observed incidence 2015-2019 | 4.05 | 4.18 | 5.11 | 4.29 | 4.37 | 3.92 | 3.51 | 4.26 |
| Expected incidence (95% credible intervals) | 3.51 (2.10-5.34) | 3.79 (2.25-5.62) | 4.36 (2.67-6.47) | 3.79 (2.25-5.62) | 4.08 (2.53-6.04) | 3.51 (1.97-5.20) | 3.09 (1.83-4.78) | 3.65 (2.11-5.48) |
| Relative incidence rate ratio (95% credible intervals) | 0.88 (0.58-1.47) | 0.67 (0.45-1.12) | 0.97 (0.65-1.58) | 1.07 (0.72-1.81) | 1.00 (0.67-1.61) | 0.92 (0.62-1.64) | 1.00 (0.65-1.69) | 0.81 (0.54-1.40) - |
| Vaccine doses per population | 0.04 | 0.06 | 0.08 | 0.14 | 0.26 | 0.29 | 0.17 | 0.07 |

Incidence is given as the number of biopsy-proven glomerulonephritis cases (IgAN, PINGN, MCD) per million population aged >18 years. Vaccine doses per population are given as doses per population. The expected incidence was calculated using a Bayesian model based on each month for the years 2015-2019. The relative incidence rate denotes the ratio between observed and expected cases.

**Supplementary Table 5. Estimated risk ratio for the development of biopsy-proven glomerulonephritis (excluding membranous nephropathy) and for the development of new symptoms or laboratory abnormalities excluding patients with membranous nephropathy.**

| **Diagnosis** | **Risk ratio for biopsy proven glomerulonephritis** | **Risk ratio for symptoms or laboratory signs of glomerulonephritis** |
|---|---|---|
| Any GN, excluding MN | 0.94 (95% CI 0.62–1.42), *P*=0.85 | 1.01 (95%CI 0.60–1.70), *P*=0.99 |

Abbreviations: MN, membranous nephropathy; GN, glomerulonephritis; CI, confidence interval

**Supplementary Figure 1. Monthly incidence of IgAN, PINGN, MCD and MN from January 2015 – August 2021.**

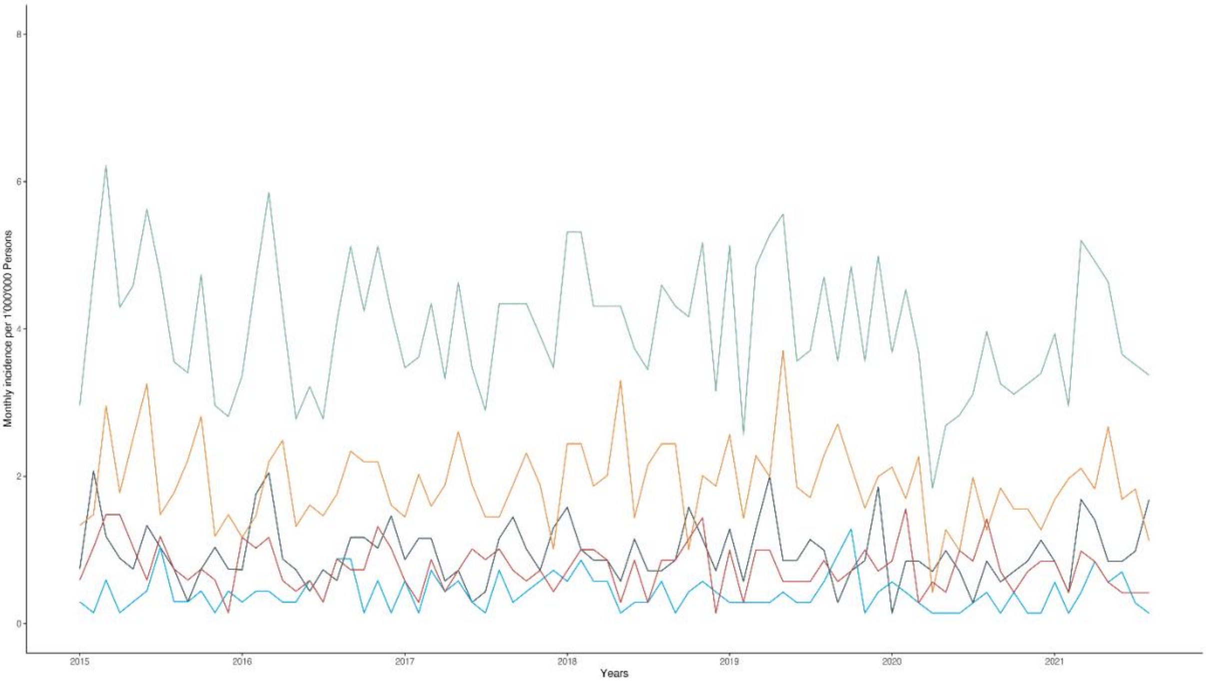


Cases of IgAN are shown in yellow, PINGN in grey, MCD in blue, MN in red and the total of all four in green.

**Supplementary Figure 2. Observed and expected incidence of glomerulonephritis during 2020, the first pandemic year.**

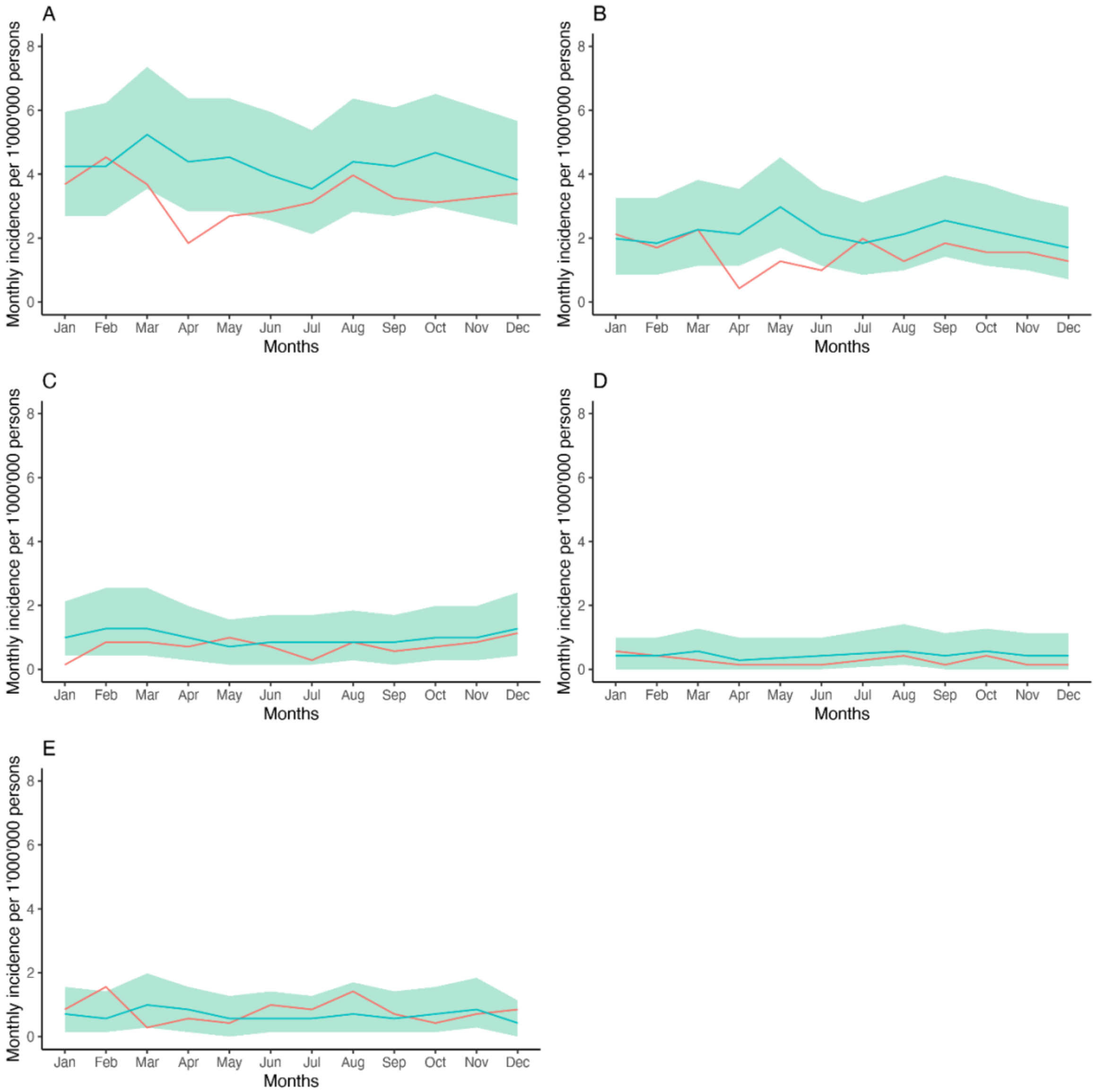


Shown is the expected incidence of glomerulonephritis for the year 2020 based on the years 2015 – 2019 (blue line) with 95% prediction intervals (green shading) as well as the observed cases during the year 2020 (red line) for the sum of all glomerulonephritis types (Panel A), IgAN (Panel B), PINGN (Panel C), MCD (Panel D) and MN (Panel E).

**Supplementary Figure 3. Frequency and timing of vaccination by dose number in patients with glomerulonephritis compared to matched controls.**

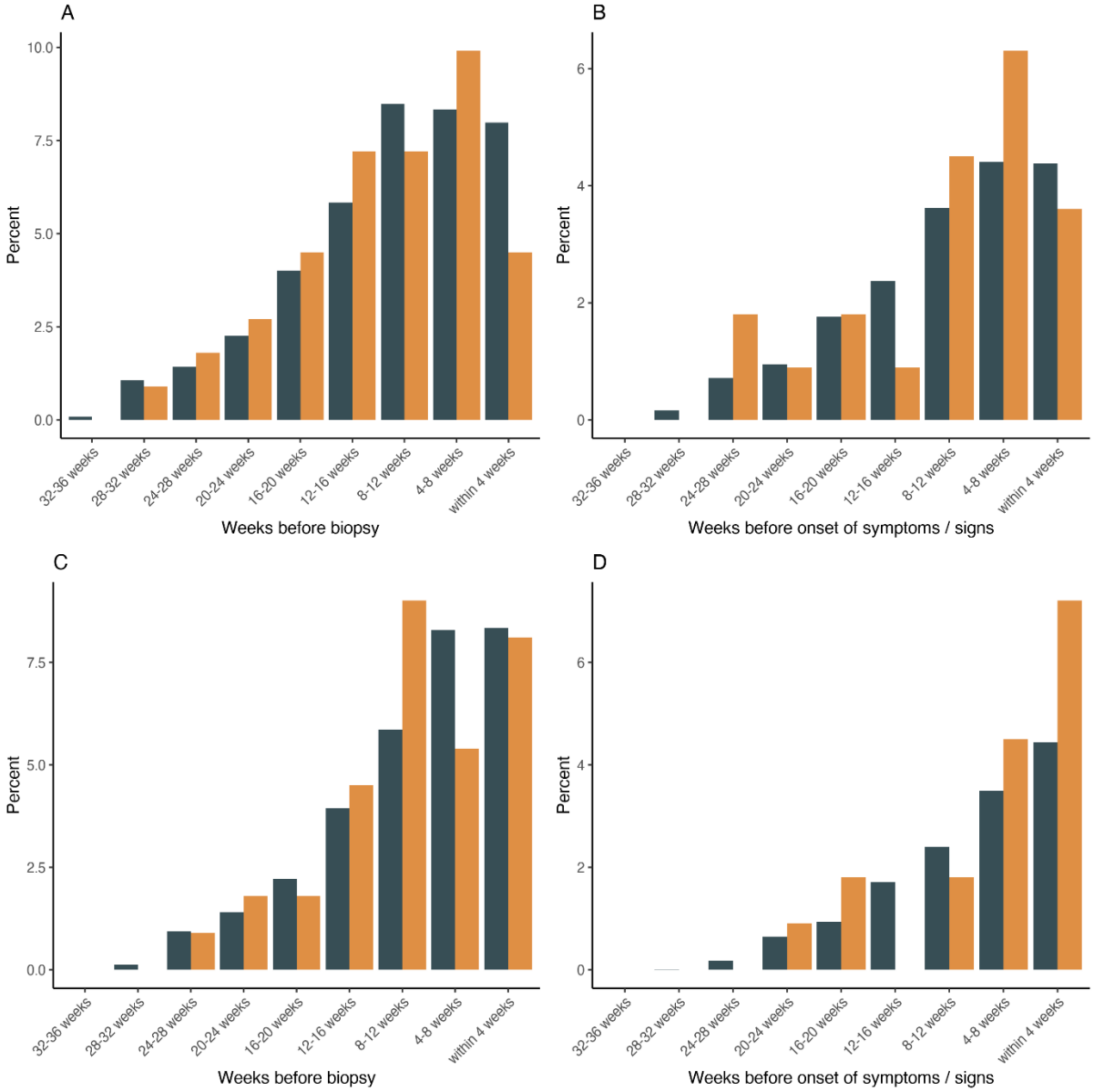


Shown in yellow is the percentage of patients with a new diagnosis of IgAN, PINGN, MCD or MN during the study period, who have received the first dose (A, B), or the second dose (C, D) during each four-week interval before renal biopsy (A, C) or onset of symptoms or detection of laboratory anomalies attributable to the renal disease or an extrarenal manifestation thereof (B, D). For comparison, the percentage of persons from a control population matched for age and timepoint during the vaccination campaign are shown in grey.

**Supplementary Figure 4. Observed and expected incidence of glomerulonephritis during the vaccination campaign, excluding membranous nephropathy.**

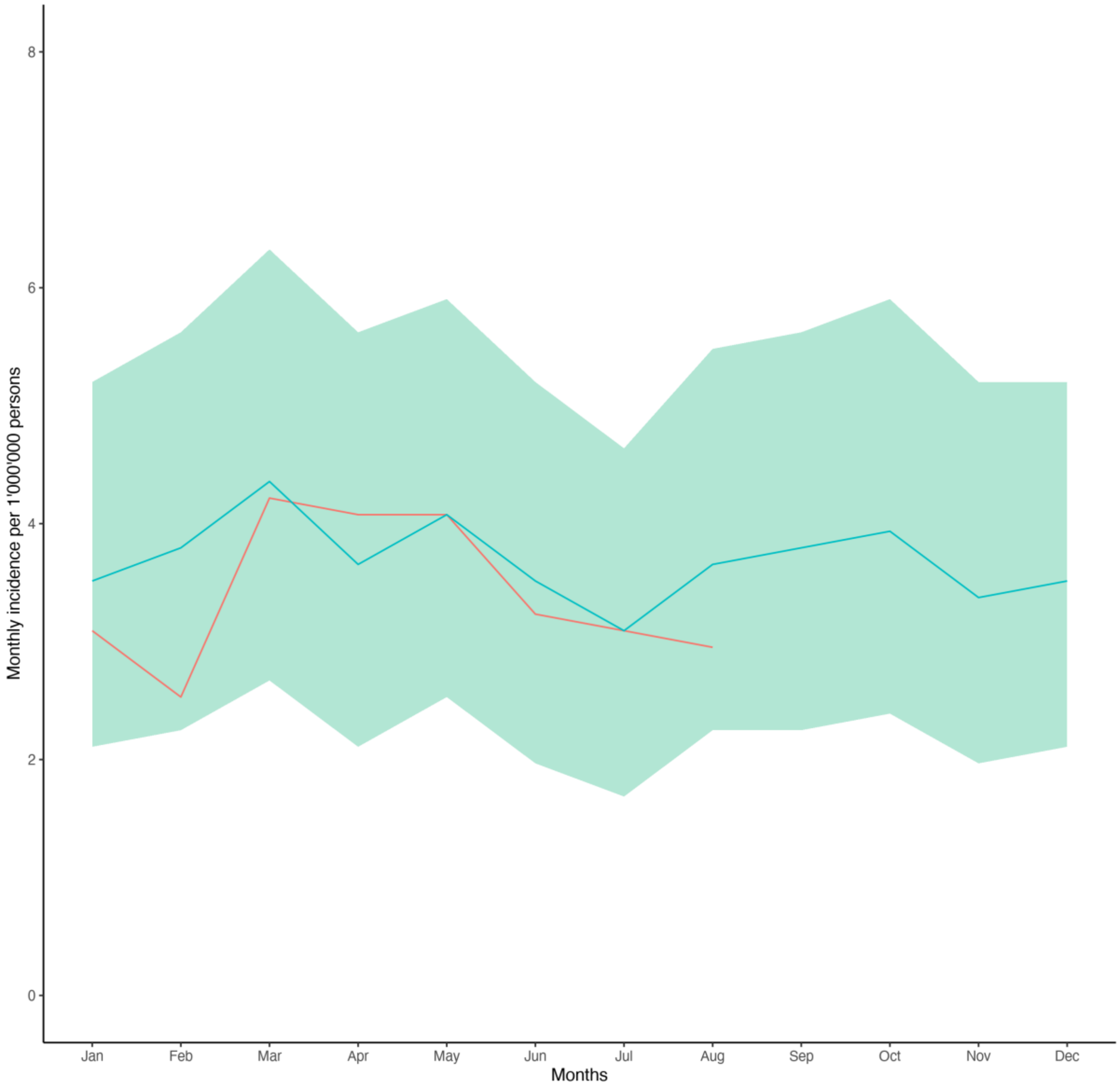


Shown is the expected incidence of glomerulonephritis based on the years 2015 – 2019 (blue line) with 95% prediction intervals (green shading) as well as the observed cases during January – August 2021 (red line) for the sum of IgAN, PINGN and MCD.

**Supplementary Figure 5. Frequency and timing of vaccination in patients with glomerulonephritis (excluding membranous nephropathy) compared to matched controls.**

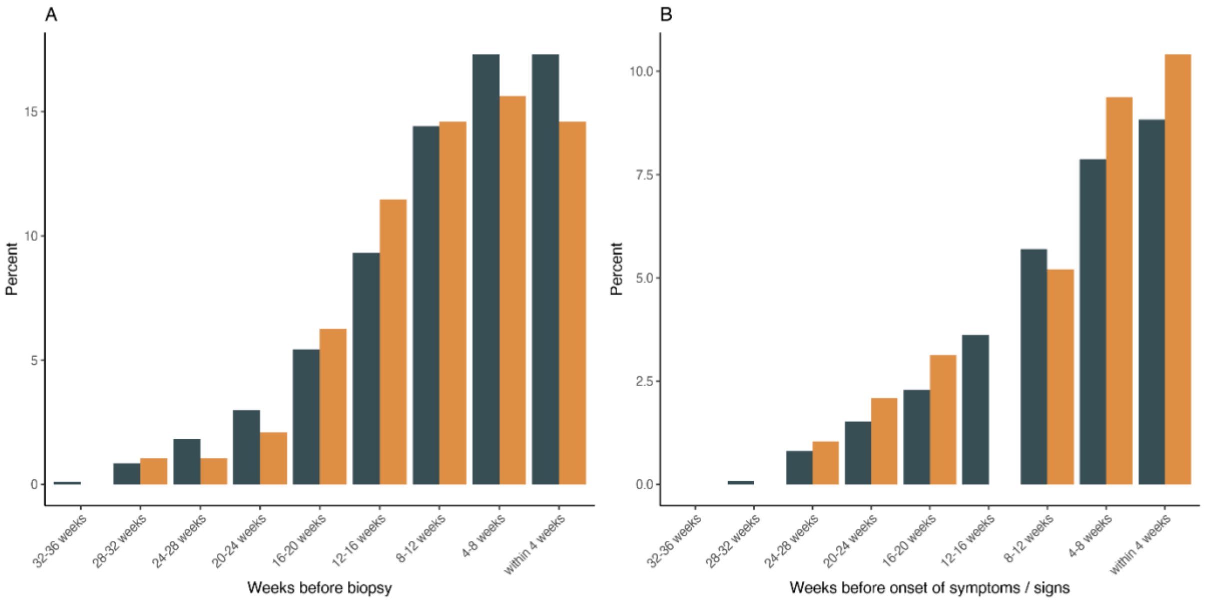


Shown in yellow is the percentage of patients with a new diagnosis of IgAN, PINGN or MCD, excluding patients with membranous glomerulonephritis during the study period, who have received any vaccine dose during each four-week interval before renal biopsy. For comparison, the percentage of persons from a control population matched for age and timepoint during the vaccination campaign are shown in grey.
